# Factors Associated with Maintenance of an Improved Ejection Fraction: An Echocardiogram-based Registry Study

**DOI:** 10.1101/2023.05.10.23289822

**Authors:** Brenna McElderry, Thomas O’Neill, Brian P. Griffin, Vidyasagar Kalahasti, Benico Barzilai, Andrei Brateanu

## Abstract

**Background:** Heart failure with improved ejection fraction (HFimpEF) is increasingly recognized as a sizable and distinct entity. While the features associated with improved ejection fraction have been explored and new guidelines have emerged, factors associated with sustaining an improved ejection fraction over time have not been defined.

**Objective:** We aimed to assess factors associated with maintenance of an improved ejection fraction in a large real-world patient cohort.

**Methods:** A total of 7,070 participants with heart failure with improved ejection fraction and a subsequent echocardiogram (ECHO) performed after at least nine months of follow-up were included in a retrospective study conducted at the Cleveland Clinic in Cleveland, OH. Multiple logistic regression models, adjusted for demographics, comorbidities, and medications were built to identify characteristics and therapeutic interventions associated with maintaining an improved ejection fraction.

**Results:** Mean age (SD) was 64.9 (13.8) years, 62.7% were men, and 75.1% were White. White race and the use of angiotensin-converting enzyme inhibitors, angiotensin-receptor blockers, or angiotensin receptor-neprilysin inhibitors were correlated with maintaining the ejection fraction at least nine months after ejection fraction improvement. In contrast, male sex, or having atrial fibrillation/flutter, coronary artery disease, history of myocardial infarction, presence of an implanted cardioverter defibrillator, and use of loop diuretics were correlated with a decline in ejection fraction after previously documented improvement.

**Conclusion:** Continued use of renin-angiotensin aldosterone system inhibitors was associated with maintaining the ejection fraction beyond the initial improvement phase.

## Introduction

Heart failure with improved ejection fraction (HFimpEF) has been increasingly recognized as a distinct clinical entity, with unique characteristics that differ from the long-established syndromes of heart failure with reduced ejection fraction (HFrEF) and heart failure with preserved ejection fraction (HFpEF).^1-7^ HFimpEF is currently defined by an initial clinical presentation of heart failure and an accompanying left ventricular ejection fraction (LVEF) ≤ 40%, with subsequent improved LVEF on follow-up imaging. While these patients can now be identified with routine echocardiography, assessing and stratifying their likelihood for maintained LVEF > 40%, as well as their risk for future cardiac events, remains a challenge.^5^ While current guidelines support continuing HFimpEF patients on the same guideline-directed medical therapy (GDMT) recommended for all HFrEF patients, evidence remains relatively limited as to which characteristics are key prognostic indicators in this heterogenous group of patients.^1^

A randomized trial aimed to assess the phased withdrawal of GDMT in patients with a history of dilated cardiomyopathy and improved ejection fraction demonstrated a higher rate of HFrEF relapse in patients that discontinue GDMT.^8^ These findings encouraged the revision of the terminology from heart failure with recovered ejection fraction (HFrecEF) to HFimpEF, since studies have demonstrated that the improved ejection fraction does not represent a true recovery of normal cardiac function. Factors associated with HFimpEF have been previously described.^9-11^ Evidence to date suggests that younger age, female sex, and the lack of history of ischemic heart disease have all been associated with initial improvement in ejection fraction.^12,13^ However, factors associated with maintenance of an improved LVEF remain largely unknown.

Using a large cohort of patients with HFrEF who demonstrated subsequent improvement in LVEF, we assessed the clinical characteristics and therapeutic interventions associated with maintaining an improved LVEF beyond nine months after the initial improvement phase.

## METHODS

### Study Population

This was a retrospective, single center cohort study of 7,070 adult patients diagnosed with HFimpEF after a previously documented HFrEF between 2010 and 2020 at the Cleveland Clinic in Cleveland, OH, USA.

The echocardiogram (ECHO) data was obtained through the Cleveland Clinic’s Heart, Vascular, Thoracic Institute Data Warehouse ECHO domain. Patients 18 years or older, with at least three transthoracic echocardiograms obtained as part of regular clinical care and available during the study follow-up, were selected. The time interval between the first and second ECHO, and the time interval between the second and third ECHO were at least 1 and 9 months, respectively (see **Figure 1** – Study Flow Diagram). The study cohort included patients with LVEF of 40% or less on the first ECHO, and LVEF greater than 40% on the second ECHO, with at least a 10% absolute increase in the LVEF from the first to the second ECHO. Exclusion criteria were cardiac transplantation, or a left ventricular assist device (LVAD) implantation between the first and second ECHO, and patients diagnosed with hypertrophic cardiomyopathy (**Figure 1**). The study protocol was approved by the Cleveland Clinic Institutional Review Board.

**Figure 1.**
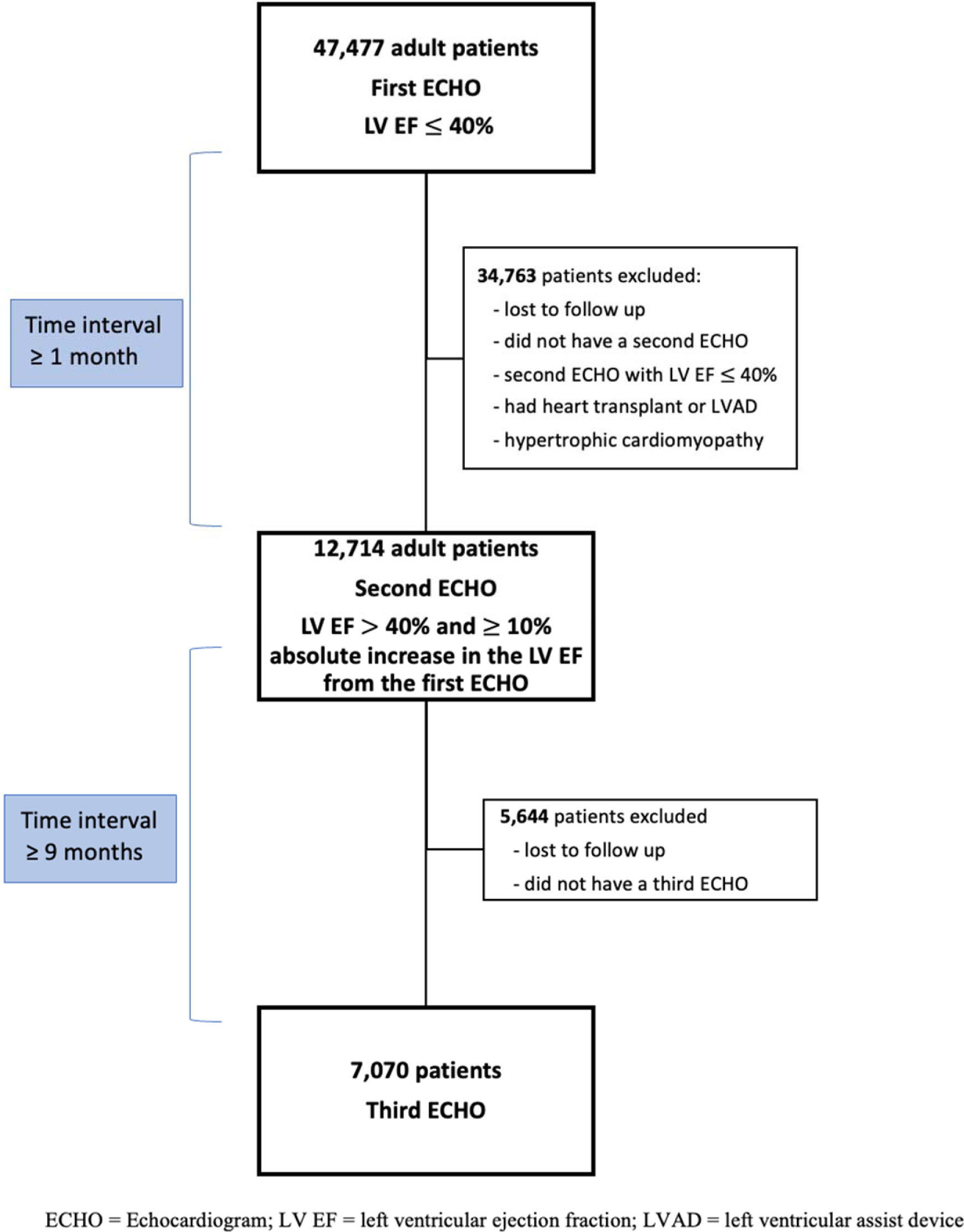
Study Flow Diagram.

### Data Collection

#### Description of Echocardiogram Parameters

Assessment of cardiac function was performed by echocardiography according to the American Society of Echocardiography guidelines.^14^ LVEF was calculated using diastolic and systolic left ventricular volumes measured by the biplane Simpson’s rule method: LVEF = ((End Diastolic Volume – End Systolic Volume)/ End Diastolic Volume)×100. These LVEF measurements were obtained from the echocardiogram reports in the Electronic Medical Record. A LVEF cutoff of ≤ 40% was diagnostic for HFrEF, while HFimpEF was LVEF greater than 40% with at least a 10% absolute increase in the LVEF from the index diagnosis of HFrEF, based on prior literature.^5^ The inclusion criteria of having a 10% absolute increase in LVEF also helps address the potential reporting errors, given the ejection fraction measured during the echocardiography is subject to a margin error of approximately 5%. All participants with a diagnosis of HFimpEF who had a subsequent ECHO after nine months or more of follow-up were included in the study.

#### Outcome

The outcome of interest was a follow up LVEF, obtained at least nine months (third ECHO) after a previously documented HFimpEF. Maintaining an improved LVEF was defined as an EF measured during the third echocardiogram (ECHO) that was equal to or higher than the EF measured during the second ECHO.

#### Covariates

Information on covariates collected at the time of the second ECHO (study baseline) was obtained from the electronic medical records, including demographic and physical measures, and medical history. History of cardiovascular disease to include coronary heart disease, myocardial infarction or revascularization, heart failure, stroke, peripheral vascular disease, chronic kidney disease, and chronic obstructive pulmonary disease, were identified using International Classification of Disease (ICD) 10 codes. Information on medications was collected via the active medication list in the patient’s electronic medical record at the time of the third ECHO. This was done in order to show continued use of GDMT and other medications after the baseline second ECHO was obtained. All medications prescribed for outpatient use were categorized into drug classes. Smoking status was categorized as current, former, or never smokers. Chronic kidney disease (CKD) was defined as an estimated glomerular filtration rate below 60 ml/min/1.73m^2^.

#### Statistical Analyses

Standard descriptive statistics were used to characterize the HFimpEF cohort at study baseline, stratified by LVEF % quartiles. Continuous variables were expressed as mean (SD) or median (interquartile range [IQR]) and compared using t tests or Wilcoxon rank-sum tests as appropriate. Categorical variables were expressed as proportions and compared using χ2 test.

Logistic regression models were used to estimate the factors associated with maintaining an improved LVEF (third ECHO), at least nine months from the echocardiographic diagnosis of HFimpEF (second ECHO). Covariates used for adjustment in the models included age, sex, race, tobacco use, history of cardiovascular disease, diabetes, chronic kidney disease, and medications (angiotensin converting enzyme inhibitor, angiotensin receptor blocker, beta-blockers, mineralocorticoid receptor antagonists, nitrates, digoxin, and loop diuretics).

All models were tested for linearity of continuous variables of interest. Given the small number of missing covariate data and the large data set, imputation was not performed; participants with missing data were excluded from the multivariable analyses. Model assumptions were tested using residual values versus fitted plots, normal Q-Q plots of standardized residuals, plots of standardized residuals versus leverage, scale-location plots, and Cook’s distance. For all analyses a two-sided p-value of 0.05 was considered statistically significant. All analyses were performed using IBM Corp. Released 2020. IBM SPSS Statistics for Windows, Version 27.0. Armonk, NY: IBM Corp.

## RESULTS

In the total cohort of 7,070 participants with HFimpEF, the mean age (SD) was 64.9 (13.8) years, 4,436 (63%) were men, 1,463 (21%) were Black, and 3,709 (53%) had a history of coronary artery disease. There were 2,819 (40%) participants with CKD.

Compared to participants in the lowest quartile, those in the highest quartile of baseline LVEF were more likely to be younger, women, with better cardiovascular profiles (less likely to have a history of coronary artery disease, myocardial infarction, coronary artery bypass grafting, percutaneous coronary intervention, cardiac resynchronization therapy, or an implantable cardioverter defibrillator). Baseline characteristics of the study population according to baseline LVEF quartiles are shown in **Table 1**.

**Table 1.** Baseline characteristics of study participants by left ventricular ejection fraction (LVEF) quartiles.

|  | Overall<br>N = 7070 | LVEF < 46<br>%<br>N = 1767 | LVEF 46-50<br>%<br>N = 1985 | LVEF 50-55<br>%<br>N = 1565 | LVEF > 55<br>%<br>N = 1753 | P Value |
| --- | --- | --- | --- | --- | --- | --- |
| Age, y | 64.9 ± 13.8 | 65.4 ± 13.6 | 65.3 ± 13.9 | 65.0 ± 13.6 | 63.9 ± 14.1 | 0.004 |
| Men | 4436 (62.7) | 1175 (66.5) | 1302 (65.6) | 965 (61.7) | 994 (56.7) | <0.001 |
| Race |  |  |  |  |  |  |
| White | 5312 (75.1) | 1360 (77) | 1487 (74.9) | 1184 (75.7) | 1281 (73.1) | 0.035 |
| Black | 1463 (20.7) | 345 (19.5) | 397 (20.0) | 322 (20.6) | 399 (22.8) |  |
| BMI | 29.5 ± 7.0 | 29.7 ± 6.7 | 29.5 ± 6.8 | 29.7 ± 7.0 | 29.1 ± 7.4 | 0.040 |
| Current smoking | 601 (8.5) | 161 (9.1) | 148 (7.5) | 153 (9.8) | 139 (7.9) | 0.023 |
| Hypertension | 5819 (82.3) | 1432 (81.0) | 1600 (80.6) | 1280 (81.8) | 1507 (86.0) | <0.001 |
| Hyperlipidemia | 4994 (70.6) | 1255 (71.0) | 1401 (70.6) | 1075 (68.7) | 1263 (72.0) | 0.197 |
| Diabetes mellitus | 2675 (37.8) | 681 (38.5) | 751 (37.8) | 579 (37.0) | 664 (37.9) | 0.839 |
| CKD | 2819 (39.9) | 682 (38.6) | 809 (40.8) | 600 (38.3) | 728 (41.5) | 0.146 |
| COPD | 2032 (28.7) | 500 (28.3) | 566 (28.5) | 471 (30.1) | 495 (28.2) | 0.606 |
| Atrial<br>fibrillation/flutter | 4045 (57.2) | 954 (54.0) | 1176 (59.2) | 928 (59.3) | 987 (56.3) | 0.003 |
| Coronary artery<br>disease | 3709 (52.5) | 1021 (57.8) | 1083 (54.6) | 802 (51.2) | 803 (45.8) | <0.001 |
| History of MI | 1888 (26.7) | 507 (28.7) | 557 (28.1) | 380 (24.3) | 444 (25.3) | 0.008 |
| History of CABG | 1248 (17.7) | 356 (20.1) | 374 (18.8) | 253 (16.2) | 265 (15.1) | <0.001 |
| History of PCI | 3017 (42.7) | 765 (43.3) | 852 (42.9) | 664 (42.4) | 736 (42.0) | 0.872 |
| ICD | 1868 (26.4) | 576 (32.6) | 545 (27.5) | 390 (24.9) | 357 (20.4) | <0.001 |
| CRT | 2071 (29.3) | 604 (34.2) | 615 (31.0) | 426 (27.2) | 426 (24.3) | <0.001 |
| ACEi, ARB, or<br>ARNI | 4556 (64.4) | 1253 (70.9) | 1373 (69.2) | 988 (63.1) | 942 (53.7) | <0.001 |
| Beta blockers | 5388 (76.2) | 1475 (83.5) | 1603 (80.8) | 1202 (76.8) | 1108 (63.2) | <0.001 |
| MRA | 1575 (22.3) | 485 (27.4) | 511 (25.7) | 343 (21.9) | 236 (13.5) | <0.001 |
| Nitrates | 572 (8.1) | 179 (10.1) | 183 (9.2) | 103 (6.6) | 107 (6.1) | <0.001 |
| Loop diuretics | 3283 (46.4) | 892 (50.5) | 994 (50.1) | 721 (46.1) | 676 (38.6) | <0.001 |
| Digoxin | 490 (6.9) | 134 (7.6) | 158 (8.0) | 89 (5.7) | 109 (6.2) | 0.023 |

ACEi = angiotensin converting enzyme inhibitors; ARB = angiotensin receptor blockers; ARNI = angiotensin receptor neprilysin inhibitors; BMI = body mass index; CABG = coronary artery
bypass graft; CKD = chronic kidney disease; COPD = chronic obstructive pulmonary disease; CRT = cardiac resynchronization therapy; ICD = implantable cardioverter defibrillator; LVEF = left ventricular ejection fraction; MI = myocardial infarction; MRA = mineralocorticoid receptor antagonist; PCI = percutaneous coronary intervention

From the total cohort, 4,608 participants had measured left ventricular volumes. Median left ventricular end diastolic volume (LVEDV) and left ventricular end systolic volume (LVESV) were 114 ml (IQR 89-145 ml) and 55 ml (IQR 40-72 ml), respectively (**Figure 2**).

**Figure 2.**
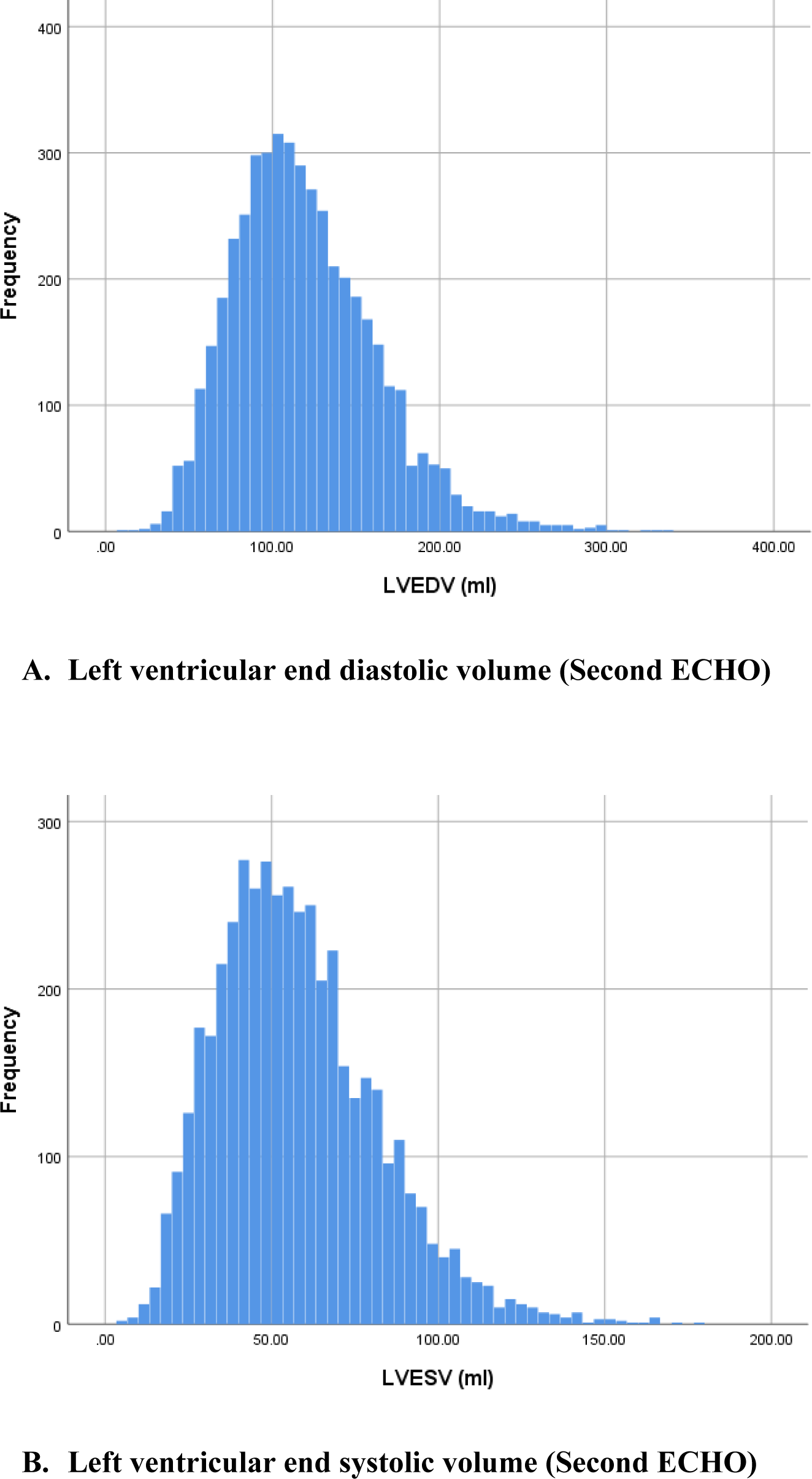
Distribution of Left Ventricular End Diastolic Volume (A) and Left Ventricular End Systolic Volume (B) in participants with HFimpEF (Second ECHO) A total of 4,608 participants had measured left ventricular volumes. Median left ventricular end diastolic volume (LVEDV) and left ventricular end systolic volume (LVESV) were 114 ml (IQR 89-145 ml) (2A) and 55 ml (IQR 40-72 ml) (2B), respectively.

### Longitudinal Follow-up of Participants with Heart Failure with Improved Ejection Fraction

The median follow-up time of participants with HFimpEF was 484 days (IQR 368-736 days). The distribution of LVEF at the third ECHO is depicted in **Figure 3**. The LVEF remained generally stable during the study period, from a median of 50% (IQR 46-55%) at the diagnosis of HFimpEF (second ECHO), to 51% (IQR 44-57%) at the follow-up echocardiogram (third ECHO). However, participants with HFimpEF in the lower LVEF quartile at study baseline (second ECHO) were less likely to maintain an LVEF more than 40% at follow-up, when compared to participants in the higher LVEF quartiles (**Figure 4**). Similarly, participants with HFimpEF in the higher LVEDV or LVESV quartile at the study baseline (second ECHO) were less likely to maintain an LVEF more than 40% at follow-up, when compared to participants in the lower LVEDV and LVESVquartiles, respectively (**Figure 5**).

**Figure 3.**
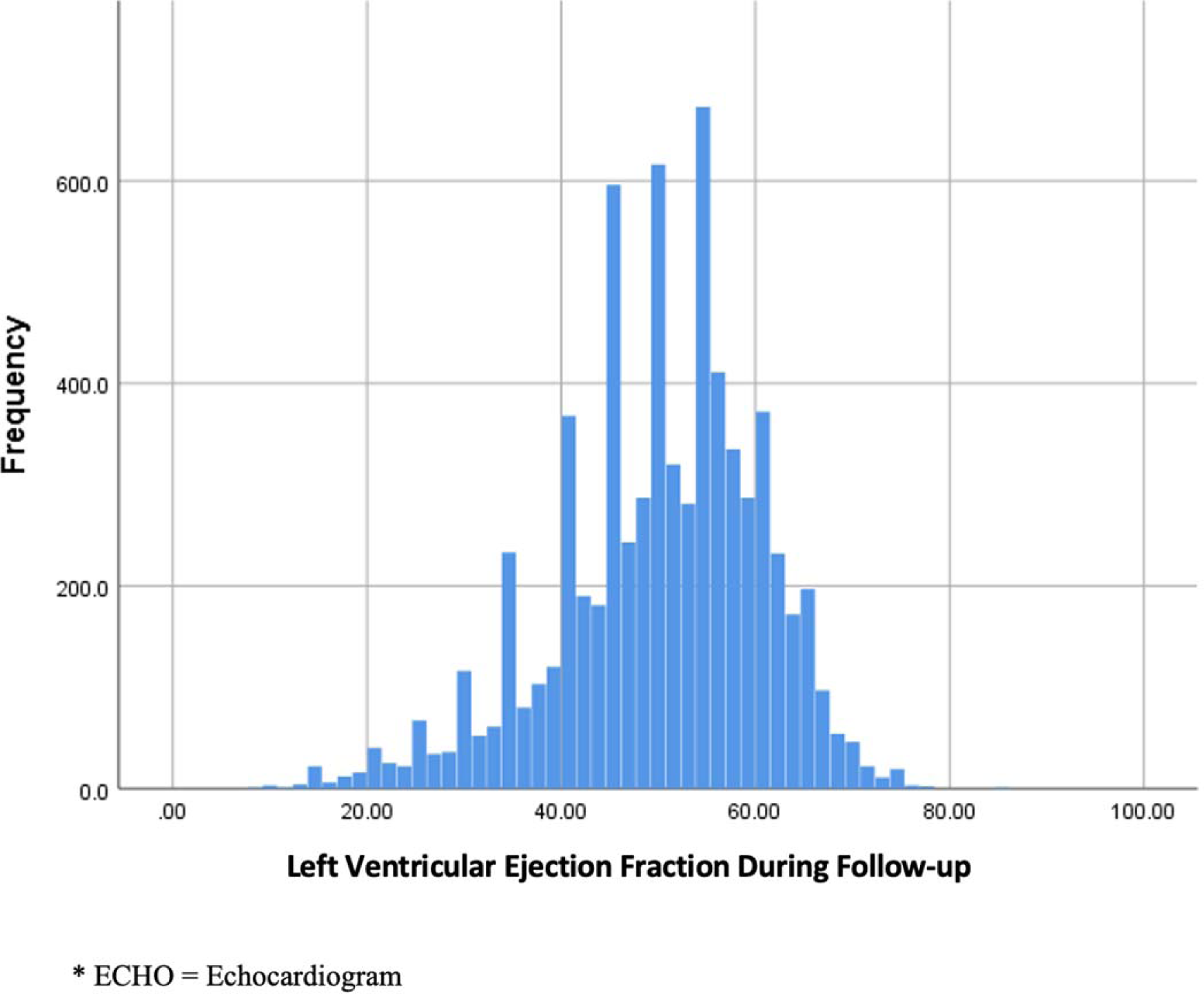
Distribution of Left Ventricular Ejection Fraction During Follow-up in Participants with Heart Failure with Improved Ejection Fraction (Third ECHO*) Participants with HFimpEF who were followed for nine months or more, maintained a median left ventricular ejection fraction of 51% (Interquartile Range 44-57%) at the follow-up echocardiogram.

**Figure 4.**
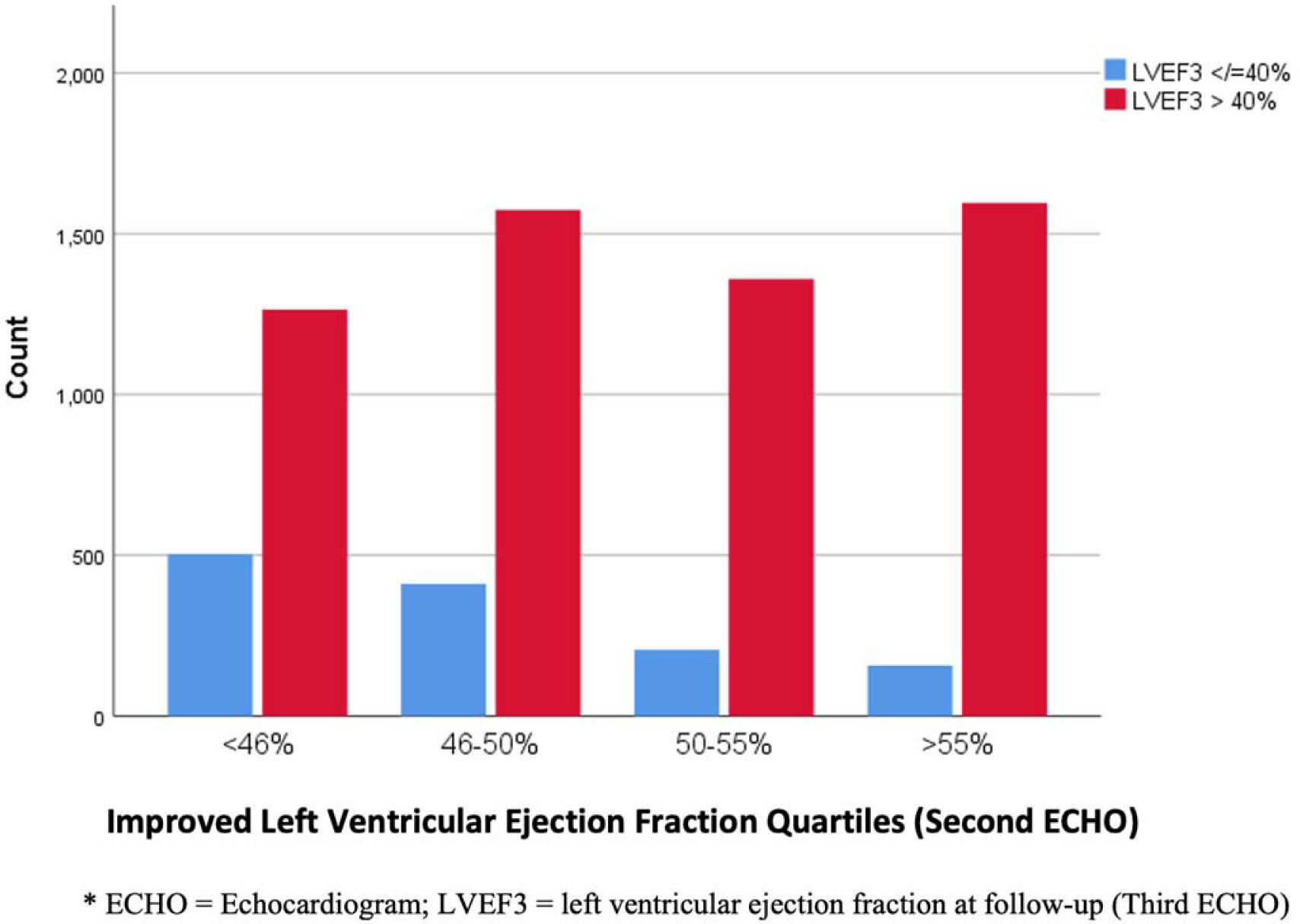
Distribution of Ejection Fraction During Follow-up (Third ECHO) by Baseline Left Ventricular Ejection Fraction Quartiles (Second ECHO) in Participants with HFimpEF. Participants with HFimpEF in the lower LVEF quartile at study baseline (second ECHO) were less likely to maintain an LVEF more than 40% at follow-up, when compared to participants in the higher LVEF quartiles.

**Figure 5.**
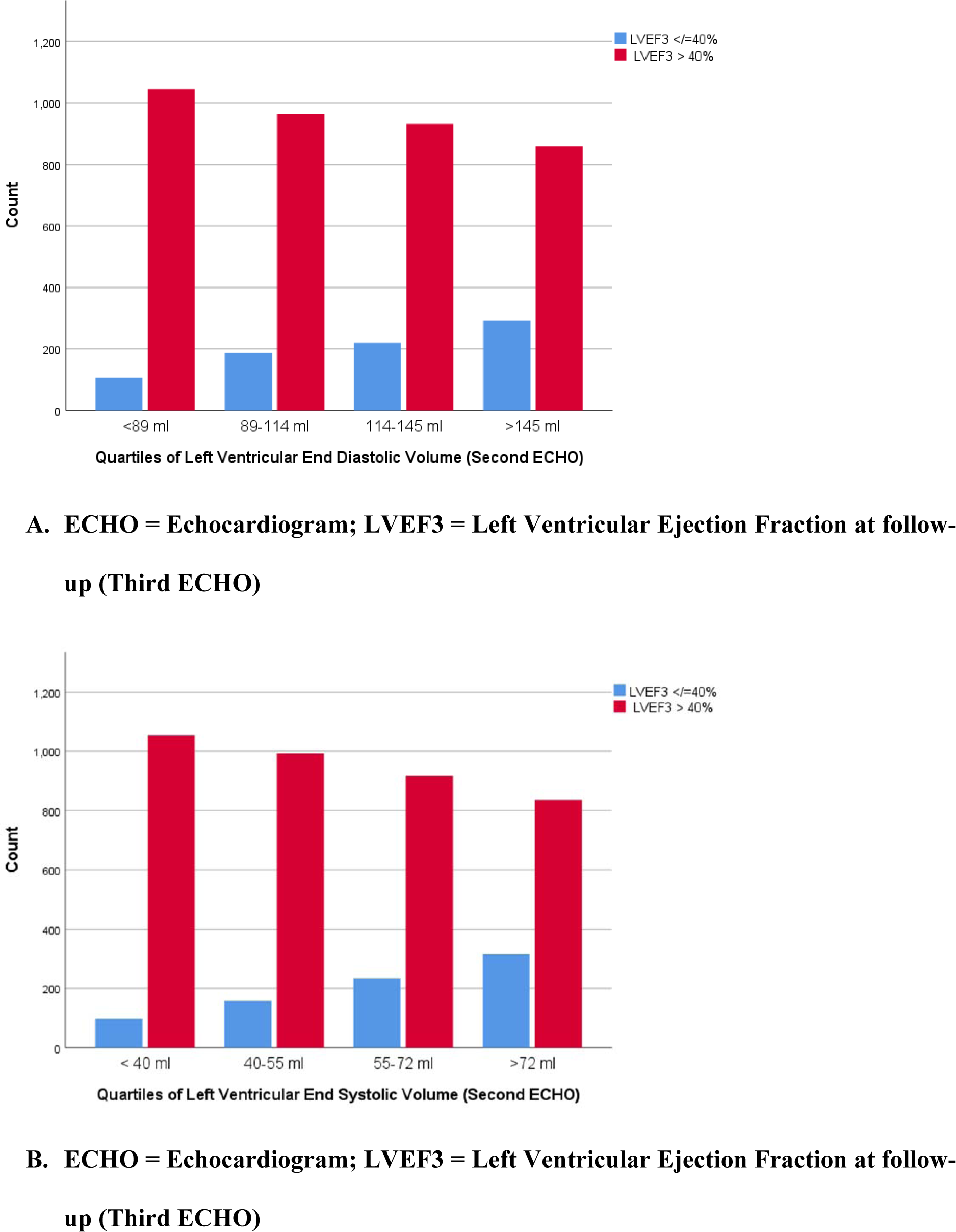
Distribution of Ejection Fraction During Follow-up (Third ECHO) by Baseline Left Ventricular End Diastolic Volume Quartiles (A) and Left Ventricular End Systolic Volume Quartiles (B) in Participants with HFimpEF (Second ECHO) Participants with HFimpEF in the higher LVEDV or LVESV quartile at the study baseline (second ECHO) were less likely to maintain an LVEF more than 40% at follow-up, when compared to participants in the lower LVEDV and LVESVquartiles, respectively.

### Factors Associated with Maintaining an Improved Left Ventricular Ejection Fraction

Characteristics associated with a greater likelihood of maintaining an improved LVEF, after adjustment for multiple covariates, included White race (Odd Ratio (OR): 1.23; 95% Confidence Interval (CI): 1.13-1.35) and continued use of angiotensin-converting enzyme inhibitors, angiotensin-receptor blockers, or angiotensin receptor-neprilysin inhibitor (OR: 1.14; 95%CI: 1.03-1.26). In contrast, male sex (OR: 0.84; 95%CI: 0.76-0.93), atrial fibrillation/flutter (OR: 0.85; 95%CI: 0.77-0.93), history of coronary artery disease (OR: 0.80; 95%CI: 0.72-0.89), history of myocardial infarction (OR: 0.75; 95%CI: 0.67-0.85), the presence of an implanted cardioverter defibrillator (OR: 0.68; 95%CI: 0.61-0.76), and use of loop diuretics (OR: 0.79; 95%CI: 0.72-0.87) were correlated with a decline in LVEF overtime in individuals with HFimpEF (**Table 2**).

**Table 2.**
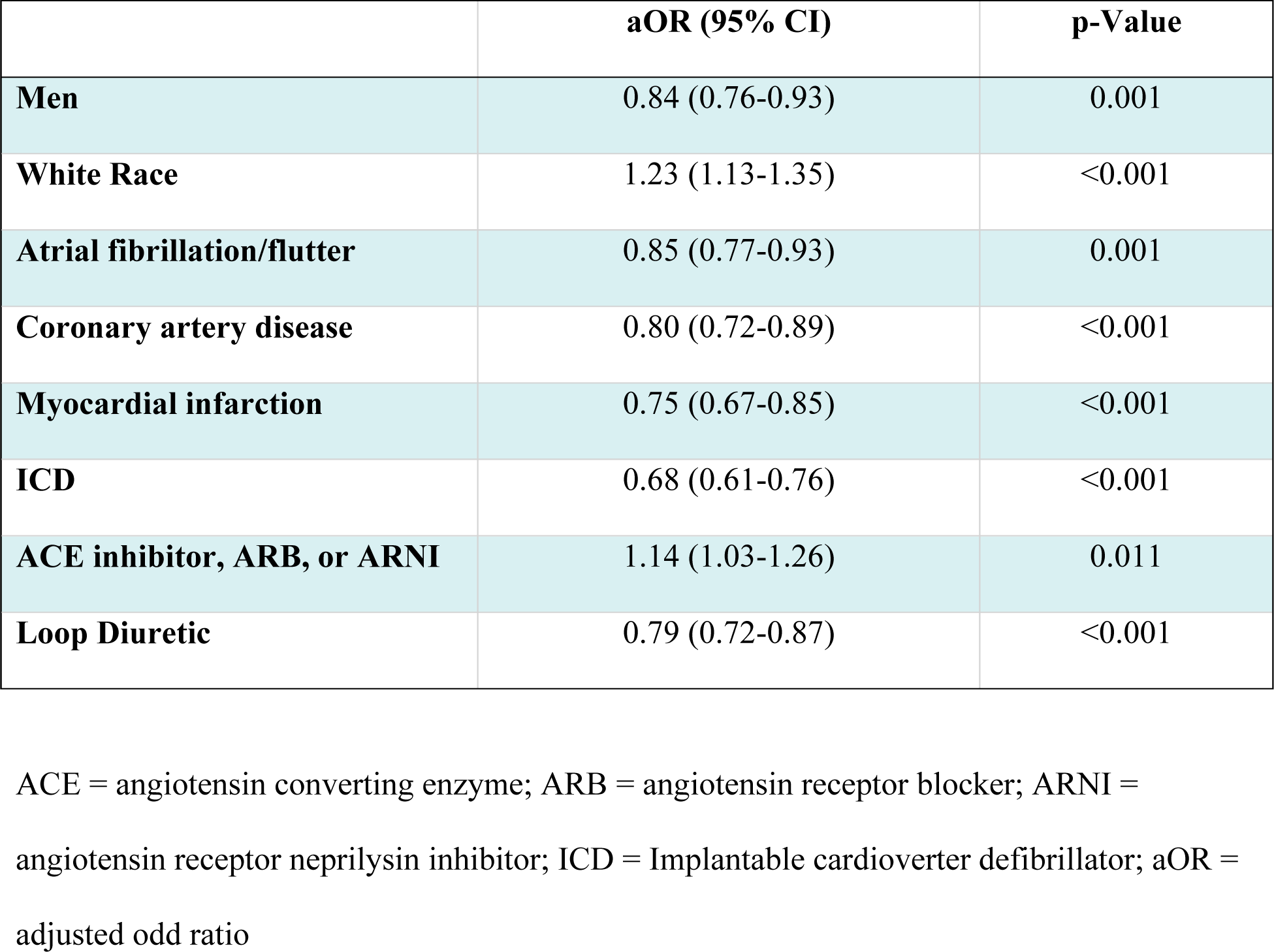
Multivariable analysis of baseline patient characteristics and medications use associated with maintaining an improved LVEF.

|  | **aOR (95% CI)** | **p-Value** |
| --- | --- | --- |
| **Men** | 0.84 (0.76-0.93) | 0.001 |
| **White Race** | 1.23 (1.13-1.35) | <0.001 |
| **Atrial fibrillation/flutter** | 0.85 (0.77-0.93) | 0.001 |
| **Coronary artery disease** | 0.80 (0.72-0.89) | <0.001 |
| **Myocardial infarction** | 0.75 (0.67-0.85) | <0.001 |
| **ICD** | 0.68 (0.61-0.76) | <0.001 |
| **ACE inhibitor, ARB, or ARNI** | 1.14 (1.03-1.26) | 0.011 |
| **Loop Diuretic** | 0.79 (0.72-0.87) | <0.001 |

ACE = angiotensin converting enzyme; ARB = angiotensin receptor blocker; ARNI = angiotensin receptor neprilysin inhibitor; ICD = Implantable cardioverter defibrillator; aOR = adjusted odd ratio

## DISCUSSION

In this large retrospective, single-center cohort study, White race and continued use of angiotensin-converting enzyme inhibitors, angiotensin-receptor blockers, or angiotensin receptor-neprilysin inhibitors were associated with a high likelihood of maintaining an improved LVEF, more than nine months after the initial diagnosis of HFimpEF. Male gender, non-White race, a diagnosis of atrial fibrillation, coronary artery disease, as well as a history of a myocardial infarction and implantable cardioverter defibrillator were factors associated with an increased risk of a decline of the LVEF in individuals with a previous diagnosis of HFimpEF.

As the etiologies and molecular basis of HFrEF have become better understood, therapeutic regimens have matured, and now allow for the possibility of improved cardiac functioning despite a HFrEF diagnosis.^15^ The cellular mechanism by which this occurs is known as left ventricular reverse remodeling, in which the heart muscle regains some of the mechanical integrity that may have previously been damaged.^16,17^ Despite these improvements in functioning, however, both basic and clinical research suggests that these improvements are not permanent, and therefore do not indicate complete normalization of the underlying pathology.^18^ Despite the accumulating evidence identifying characteristics of patients who will see improvement in their ejection fraction, few studies have assessed the factors associated with sustaining an improved ejection fraction over time. Recent research has identified echocardiographic findings, specifically global longitudinal strain, which can be used to identify subgroups of patients who may be more likely to revert back to HFrEF after initial improvement.^19^ Other studies have suggested that specific medications or medication classes may be associated with sustaining an improved ejection.^20,21^. Our single-center retrospective cohort study, which evaluated 7,070 patients, fills current gaps in knowledge, by identifying demographic information, medications use, comorbidities, and procedural interventions that are associated with maintaining an improved ejection fraction over time. Additionally, the present study adds to the body of evidence identifying therapies most important to continue, in order to maintain an improved ejection fraction, particularly in patients who may develop contraindications to GDMT medications in their lifetime in which additional therapeutic interventions are required.

Of the GDMT, Angiotensin Converting Enzyme Inhibitors, Angiotensin Receptor Blockers or Angiotensin Receptor-Neprilysin Inhibitors (ACEi/ARB/ARNI) were the only medications correlated with maintaining an improved ejection fraction in our study. When examining additional medications typically used in the setting of heart failure, the need for loop diuretics was correlated with a drop in ejection fraction in the HFimpEF population. This may be related to residual confounding, as the continuous use of diuretics may be related to the presence of other comorbidities (e.g. CKD) that may increase the risk of heart failure overtime.

Medications working on the renin-angiotensin axis have well demonstrated benefits for multiple subsets of heart failure patients, including reducing heart size and increasing ejection fraction with reverse remodeling.^18,22^ Our finding of atrial fibrillation associated with a decline in the LVEF also correlates to studied pathophysiology.^23^ The tachycardia and irregularity of atrial fibrillation theoretically results in decreased diastolic filling time and thus decreased cardiac output.^23^ Heart failure may cause atrial fibrillation as well, including mechanisms such as atrial stretch, interstitial fibrosis, and dysregulation of intracellular calcium.^23^ Atrial fibrillation is used as a predictor of worsening cardiac function and is particularly shown to be detrimental to the HFimpEF population in our study.^24^

Our study has several strengths. First, prior studies required a single demonstration of improvement in ejection fraction with minimal follow up to confirm the patients continued to stay in the HFimpEF category.^6,7,9,11,12,15,18-21^ Those more likely to originally improve the left ventricular function have a growing body of research identifying the unique demographic and medical profile associated with HFimpEF.^3-13,15-21^ We assessed longitudinally if those predictive factors remained the same over time or if certain prior comorbidities may be particularly harmful after the demonstration of reverse cardiac remodeling. Similar to others, we identified specific patients characteristics associated with a HFimpEF, including sex, race, and indicators of ischemic damage.^6-12,15,18^ Adding to the evidence, we identified the presence of atrial fibrillation and the use of ACEi/ARB/ARNIs as significant factors associated with maintaining the LVEF in patients with HFimpEF.^8-12,18-21^

Second, limited information exists on the effect of GDMT in this patient population.^8,18,20,21^ One randomized controlled trial in 50 patients with nonischemic HFimpEF showed improved outcomes with continuation of GDMT and has helped shaped current national clinical guidelines recommendations.^8^ Other studies have shown beta blockers and spironolactone were associated with improved outcomes in this population.^20,21^ Our study did not find beta blockers to be associated with maintaining the LVEF in patients with HFimpEF. One possible explanation is that, over time, patients with HFpEF may experience a decrease in sympathetic activation. Consequently, beta blockers may lose their beneficial effects in a similar manner to what has been observed in patients after a myocardial infarction.^25^

We did show that ACEi/ARB/ANRI treatment should be considered a priority and continued in patients with HFimpEF, as demonstrated in the TRED-HF trial.^8^ This becomes important when patients develop complications such as hypotension and acute kidney injury while on GDMT for HFimpEF. If a patient were to require discontinuation of such medications, quantifying the benefit of certain drug classes, as this study has done, can tailor long term treatment plans, including re-challenging with the introduction of the GDMT medications once the acute complication subsides. It also reaffirms the notion that while cardiac remodeling can occur with the use of these medications, the risk to relapse is a more realistic outcome than full recovery of the myocardium, especially in patients who have stopped GDMT.^8,16,17^ Third, our study including over 7,000 participants with HFimpEF is the largest to date, which strengthens the significance of our results.^6,7,11-13,15,19-21^ Our study has few limitations. This was a retrospective single center study, and we relied on accurate documentation of covariates and echocardiogram reports as available in the Electronic Medical Record system and the Echocardiography database. Whether the results would apply to different populations is unknown. Major interventions including heart transplantation and LVAD implantation were excluded but valvular surgeries were not excluded, however it is not expected that a valvular surgery would cause reversal in the initial improvement in left ventricular ejection fraction.

In summary, our study identified characteristics and medications correlated with maintaining an improved ejection fraction for an extended period of time. Continued use of renin-angiotensin aldosterone system inhibitors was associated with maintaining of an improved ejection fraction beyond the immediate period of improvement. Future studies are needed to assess whether the addition of newer heart failure therapies such as sodium-glucose co-transporter 2 inhibitors (SGLT2i) have similar association with maintaining an improved ejection fraction.

## Data Availability

To ensure participant privacy, the raw data used in this study are not publicly available. However, upon request, the raw data can be made accessible to authorized individuals through the Cleveland Clinic Research Institute, subject to appropriate privacy and data protection protocols.

## Non-standard Abbreviations and Acronyms

ECHO: echocardiogram
GDMT: guideline-directed medical therapy
HFimpEF: heart failure with improved ejection fraction
HFpEF: heart failure with preserved ejection fraction
HFrEF: heart failure with reduced ejection fraction
LVEF: left ventricular ejection fraction

## Acknowledgements

We are grateful to Ms. Elizabeth Hejny who facilitated echocardiographic data extraction.

